# An evaluation of barrier-strategy matching for a cervical cancer prevention intervention in Kenya: A convergent mixed methods study

**DOI:** 10.1101/2025.09.01.25334878

**Authors:** Harriet Fridah Adhiambo, Sharon Cherotich Mutai, Megan Coe, Valary Ihaji, Mary Bernadette Kerubo, Alex Kinyua, Sarah Njoroge, Lynda Myra Oluoch, Thomas A. Odeny, Michelle B Shin, Bryan Weiner, Kenneth Ngure, Katherine K. Thomas, Nelly Mugo, Sarah Gimbel

## Abstract

Implementation strategies that address context-specific barriers are critical to improving implementation success and health outcomes. However, strategies are often misaligned with the barriers they are intended to address, limiting their impact. This study evaluates the extent to which deployed implementation strategies in a cervical cancer prevention program matched the pre-identified barriers. A mixed-methods convergent parallel design was employed between January 2024 and September 2024. Barrier-strategy match was defined as selecting implementation strategies that most effectively address specific barriers to adopting the single-visit, screen, and treat approach with thermal ablation (SV-SAT+TA).

Forty participants across ten health facilities completed a structured survey and participated in small group discussions conducted at each facility to gather perspectives on how and why barriers were successfully, partially, or not addressed. Descriptive statistics summarized the survey responses, and the framework method guided the analysis of the qualitative data.

Findings were integrated by merging the quantitative and qualitative results and using a narrative approach during interpretation and reporting. The majority (77%, n=32) of the participants were female, and 95% (n=38) were frontline healthcare providers. A large proportion (81%, n=17) of the reported pre-identified barriers were health system related. Overall, the implementation strategies were well-matched to the barriers, with 71% (n=15) reported as successfully addressed. Several strategies were deployed, with some addressing multiple barriers simultaneously. Key themes associated with successful barrier-strategy matching included streamlining and optimizing clinic processes to improve service accessibility and efficiency, capacity building to enhance provider knowledge, skills, and quality of cervical cancer prevention service delivery, and strengthening community engagement and communication to improve awareness and service uptake. Approximately 10% (n=2) of the barriers were partially addressed, while 19% (n=4) were not addressed.

Barriers partially addressed and those not addressed were primarily associated with structural problems, including staffing challenges, workforce instability, infrastructural limitations, and lack of support systems for patient follow-up. Aligning implementation strategies with context-specific barriers improves the likelihood of successful intervention delivery and strengthens the overall impact of cervical cancer prevention efforts.

## Introduction

Advances in cervical cancer prevention have significantly led to reductions in incidence and mortality, particularly in high-income countries.^1^ However, the global burden remains substantial, with low- and middle-income countries (LMICs) bearing the greatest impact.^2,3^ In 2022, approximately 350,000 cervical cancer-related deaths were reported, 94% of which occurred in LMICs primarily due to limited availability and suboptimal utilization of preventive services such as Human papillomavirus (HPV) vaccination and screening.^4–6^ Therefore, context-specific implementation strategies are needed to effectively implement these evidence-based preventive interventions to reduce global inequities in cervical cancer prevention.^7^

Implementation strategies are key in ensuring improvement in the adoption, implementation, and sustainability of evidence-based interventions (EBIs).^8,9^ However, their impact is often limited by misalignment or a mismatch between the selected strategies and the barriers they are designed to address.^10–12^ Several implementation strategies have been employed to improve screening and precancer treatment uptake in cervical cancer prevention. These include educational interventions, the establishment of mobile clinics, and financial incentives, among others.^13,14^ Similar to other interventions, these strategies are frequently selected without a systematic approach, insufficient contextual understanding, and inadequate stakeholder engagement.^15,16^ As a result, resources may be wasted on ineffective strategies, and the intervention may fail to achieve its intended outcomes. This highlights a critical gap; there is limited evidence on whether deployed implementation strategies in cervical cancer prevention interventions actually address the barriers they are intended to overcome. As Mosquera et al. recommend, assessing the fit between barriers and strategies is essential to improving implementation effectiveness.^17^

To address this gap, we evaluated whether the deployed implementation strategies addressed barriers to implementing the single visit, screen, and treat approach with thermal ablation (SV-SAT+TA) using a convergent mixed methods approach.

## Methods

### Ethics Statement

This study received approval from the Scientific and Ethics Review Unit of the Kenya Medical Research Institute (SERU-KEMRI No. 4403) in Nairobi, Kenya, as well as the Human Subjects Institutional Review Board (STUDY00014200) at the University of Washington in Seattle, USA. All participants provided written informed consent to take part in the survey and interviews.

### Study design and setting

To evaluate how well the implementation strategies address the pre-identified barriers to TIBA’s implementation in Kenya, this longitudinal sub-study utilizes a convergent parallel mixed methods design (Fig 1).^18^ The rationale for this approach is rooted in the research question that seeks to determine whether the strategies matched the pre-identified barriers (quantitative) and to explore perspectives on how and why these barriers were addressed successfully, partially, or not at all, as well as contextual factors that influenced strategy implementation (qualitative). Barrier-strategy match was defined as the selection of implementation strategies most effective in addressing specific barriers to adopting the single-visit, screen, and treat approach with thermal ablation (SV-SAT+TA). This study’s philosophical assumptions were guided by the pragmatic worldview.^19^ The quantitative and qualitative data were collected at the same time to converge both data sets to understand the research question better. Equal weight was given to each data type (QUAN+QUAL), i.e., each data type was considered equally important for answering the research question This sub-study, conducted in Kenya, is nested within the main trial that aims to develop and evaluate implementation strategies to inform nationwide scale-up of a single visit, screen-and-treat approach with visual inspection with acetic acid (VIA) and thermal ablation for sustainable cervical cancer prevention services.^20^ TIBA, a Swahili word for “treatment,” is used for this trial to represent the single-visit, screen and treat approach with thermal ablation. TIBA is a cluster-randomized stepped wedge design being conducted across 10 reproductive health clinics in Kenya across three successive waves, each with an intensive phase lasting six months, followed by 18, 12, and 6 months of maintenance phases, respectively.^20^ The Kenyan healthcare system is tiered, comprising six levels of care, from Level I (community health units, which are the smallest) to Level VI (national referral hospitals), designed to handle increasingly complex cases. TIBA is implemented across Levels III, IV, and V health facilities located in Kiambu, Murang’a, and Embu Counties, which are either managed by faith-based organizations or the Ministry of Health (MOH). The average monthly outpatient clinic attendance in Level III is approximately 2,500, in Level IV, 5,000-8,000, and in Level V, over 15,000.^21^

**Figure 1:**
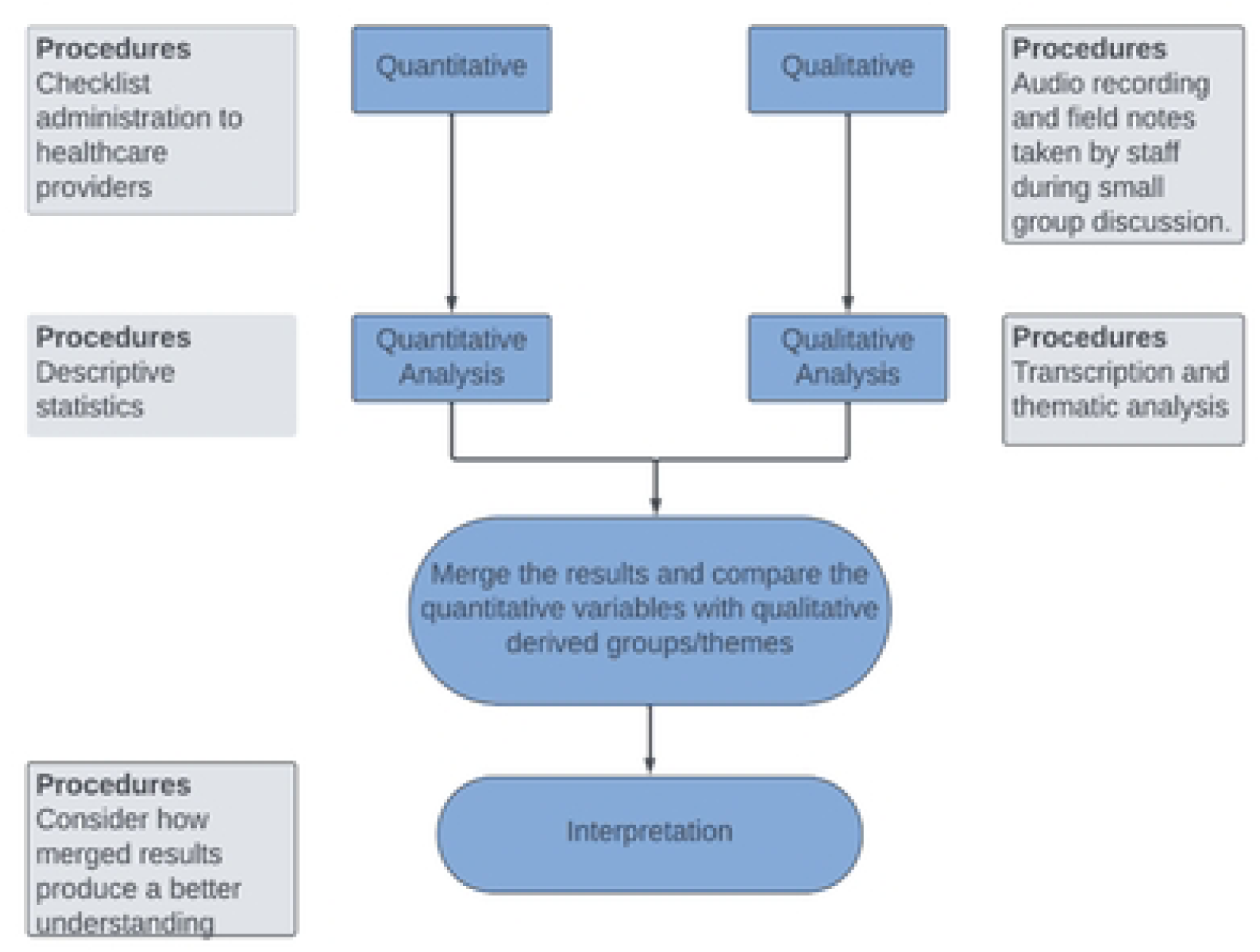
Convergent Mixed Methods Design

### Study participants

A purposive sample of health care providers offering cervical cancer prevention services at the 10 reproductive health clinics was included in the study. This sample comprised nurses, gynecologists, clinical officers, laboratory technologists, and health facility managers. Five providers, including the health facility managers, were recruited from each clinic by word of mouth and phone calls to participate in the assessment. However, the number of participants per facility ranged from 3 to 5 participants due to work-related commitments. In total, 40 participants participated in the assessment.

### Study measures and data collection procedures

We utilized a structured survey that contained two sections: one assessing the extent to which the pre-identified barriers were addressed, and the other section capturing new barriers that emerged during the implementation process. The pre-identified barriers to the implementation of TIBA were identified during a stakeholders’ meeting before the roll-out of the trial. The participants in the stakeholders’ meeting included representatives of health care providers from the health facilities, county health management teams (CHMT), policymakers from the Ministry of Health (MOH), and representatives from the Kenya National Cancer Institute (NCI). However, for this evaluation, only healthcare providers from the 10 health facilities were included, as they were directly involved in implementing the strategies.

Prior to the roll-out of TIBA, implementation strategies were matched to the barriers at the facility level. This process involved the health facility providers, in collaboration with the research team, selecting context-relevant barriers and strategies they considered feasible to implement.

The quantitative section required the participants to indicate whether each pre-identified barrier applied to their health facility and whether it was successfully, partially, or not addressed. Barriers were defined as follows:

#### Successfully addressed barriers

The barrier was initially present, but it was fully resolved or no longer hindered implementation, based on the participant’s perception.

#### Partially addressed barrier

The barrier was one that was present, and the strategy applied reduced its impact, but did not fully eliminate it, as some challenges remained.

#### Not addressed

The strategy had little or no effect, and the barrier continued to impede implementation efforts.

This approach was also applied to the section capturing emerging barriers to TIBA implementation.

Upon completion of the quantitative phase, a small group discussion was immediately co-led by a technical assistant and a qualitative research assistant, both experienced in clinical and qualitative research, respectively. The small group discussions elicited participants’ perspectives on how and why barriers were successfully, partially, or not addressed. These 50-minute discussions were conducted in English at the end of each implementation wave and were audio recorded. This data was collected between 15/01/2024 and 30/09/2024

### Analysis

#### Quantitative

Descriptive statistics were used to summarize the characteristics of participating clinics using R software, with comparisons made between government-led and faith-based facilities. Pre-identified barriers reported by participants were ranked and compared across the two facility management types. For each barrier, we reported the proportion of respondents who perceived it as successfully addressed, partially addressed, or not addressed. To determine the overall classification for the barriers, we applied the majority “rule” approach, where barriers were classified as “successfully addressed, “partially addressed,” or “not addressed” based on the response category by the majority of the respondents. This classification was then used to determine the overall barrier-strategy match by summing the number of barriers successfully, partially or not addressed across the sites. In a case where there was an equal rating between the categories, consensus on the classification was based on the review of the qualitative data and discussion between the lead author and the data collection team.

Flow diagrams were used to visually map the relationship between the reported barriers, corresponding implementation strategies, and the relevant WHO health systems Framework building blocks. This approach enabled the identification of key health systems domains targeted by the implementation strategies.

#### Qualitative

The qualitative data from each of the 10 facilities were transcribed verbatim. The framework analysis method^22,23^ was inductively applied by two authors (HAF and SCM) to identify emerging themes. This process consisted of seven stages: transcription, familiarization with the data, coding, developing an analytical framework, applying the analytical framework, charting the data into the framework matrix, and interpretation. During the familiarization stage, the authors reviewed one transcript jointly and developed the analytic framework after discussing and agreeing on the codes. The remaining nine transcripts were divided between the two authors, who applied the subsequent stages of the framework analysis, including charting the data into the matrix and interpreting the results.

#### Integration of quantitative and qualitative data

Integration involved merging the results from both the quantitative and qualitative data after separate analysis, allowing for a comparison that provides a more comprehensive understanding than either the quantitative or qualitative results alone. At the interpretation and reporting stage, integration was achieved through a narrative approach. Where applicable, we will highlight any differences that may have emerged across various management types (government-led vs faith-based) to provide contextual nuance.

## Results

We collected survey responses and conducted interviews with forty participants across 10 health care facilities. The majority of the health facilities (70%, n=7) were publicly managed. Among the forty participants, 77% were female (n=31), 95% (n=38) were frontline health care workers, with nurses representing the majority (78%, n=31) of healthcare providers offering cervical cancer preventive services. Table 1 provides a summary of the participant and clinic characteristics.

**Table 1:** Participant demographics and clinic characteristics

| Characteristics | All<br>(n, %) | Public/Government<br>(n, %) | Faith-based<br>(n, %) |
| --- | --- | --- | --- |
| No. of health facilities | 10 | 7 | 3 |
| Location |  |  |  |
| Urban | 8 (80%) | 6 (86%) | 2 (67%) |
| Rural | 2 (20%) | 1 (14%) | 1 (33%) |
| No. of participants | 40 | 29 | 11 |
| Sex |  |  |  |
| Male | 9 (23%) | 4 (14%) | 5 (45%) |
| Female | 31 (77%) | 25 (86%) | 6 (55%) |
| Cadres |  |  |  |
| Nurses | 31 (77%) | 24 (80%) | 7(70%) |
| Clinical Officers | 6 (15%) | 5(17%) | 1(10%) |
| Medical Officers/Gynecologists | 1(3%) | 0 | 1(10%) |
| Laboratory Technologists | 2 (5%) | 1(3%) | 1(10%) |
| Participant Roles |  |  |  |
| Frontline healthcare worker | 38 (95%) | 28(97%) | 10 (91%) |
| Health facility Manager | 2(5%) | 1(3%) | 1(9%) |

### Quantitative Results

During the stakeholders’ workshop, a total of 21 barriers were pre-identified by participants as factors that would negatively influence the implementation of TIBA (S1 Table). The majority of the barriers (81%, n=17) were health system-related, reported across multiple facilities. The most frequently reported barriers included staff shortages, identified by 93% of the respondents, and lack of essential commodities for screening and treatment of precancerous lesions, also reported by 93% of the respondents. These two barriers ranked as the top two implementation challenges.

Among patient-related barriers, low participant turnout (85%) and negative attitude towards cervical cancer screening (78%) were commonly reported. These were mainly driven by beliefs and fears surrounding the screening process, and ranked third and fifth, respectively (Fig 2).

**Figure 2:**
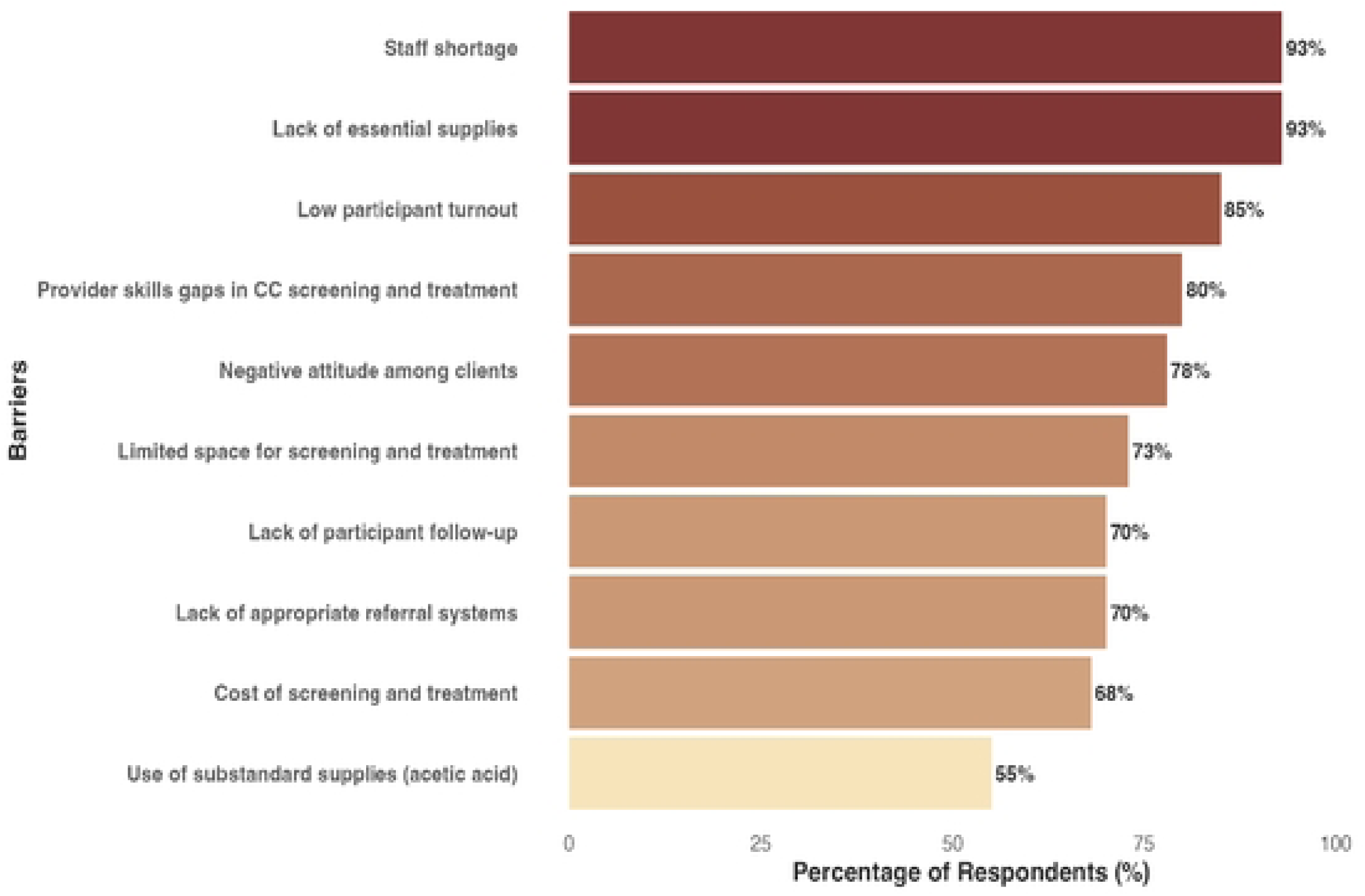
Ranking of the top ten barriers identified across healthcare facilities.

There were no differences in the barriers observed when comparing the two health facility levels, i.e., Level IV and V. However, when comparing barriers across facilities with different management levels (Fig 3), the cost of cervical cancer screening and treatment, as well as low participant turnout, were commonly reported in faith-based health facilities compared to public health facilities. The higher cost of cervical cancer preventive services in faith-based health facilities led to a lower participant turnout, as most of the women could not afford these services.

**Figure 3:**
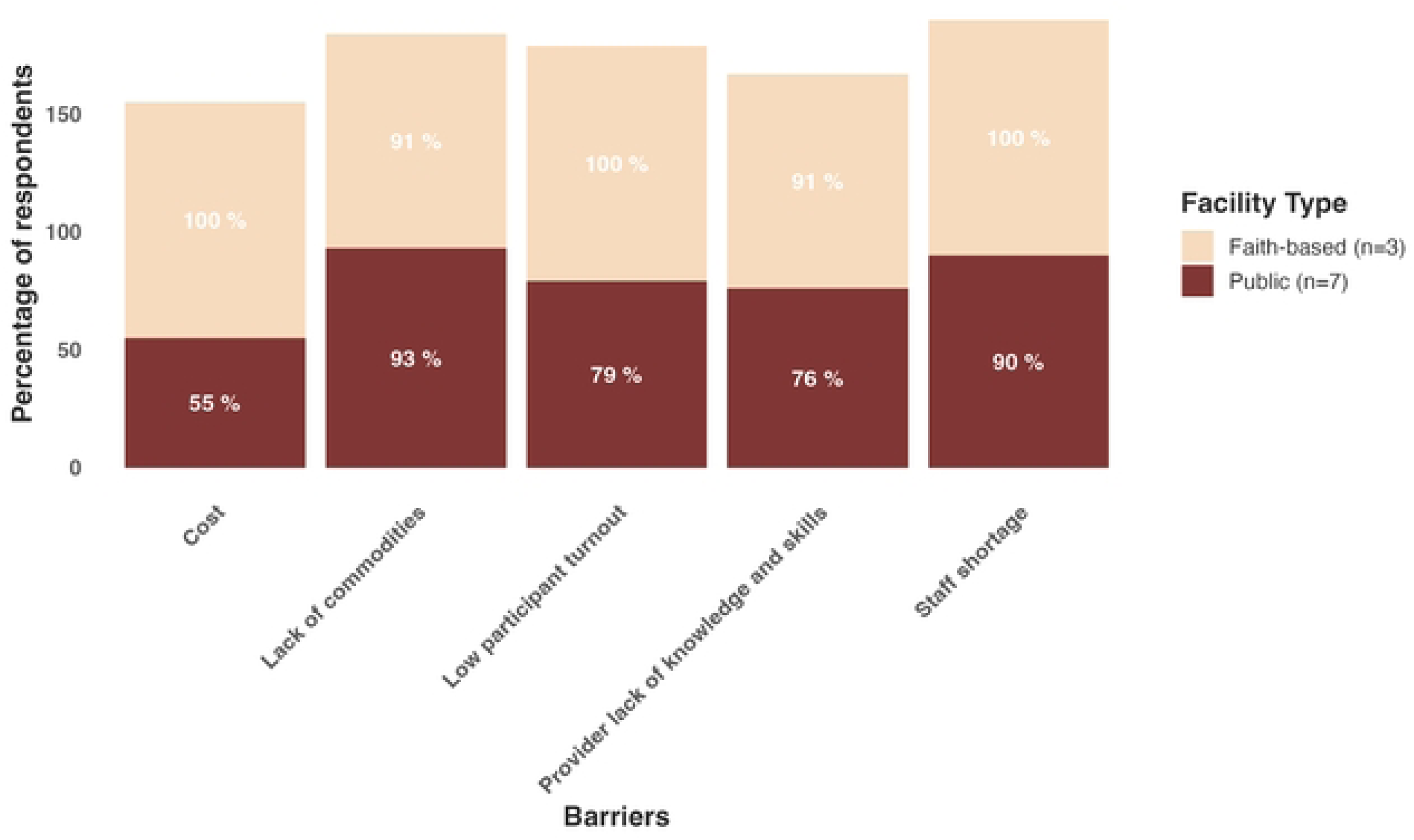
Comparison of barriers between faith-based and public health facilities

**Figure 3A.**
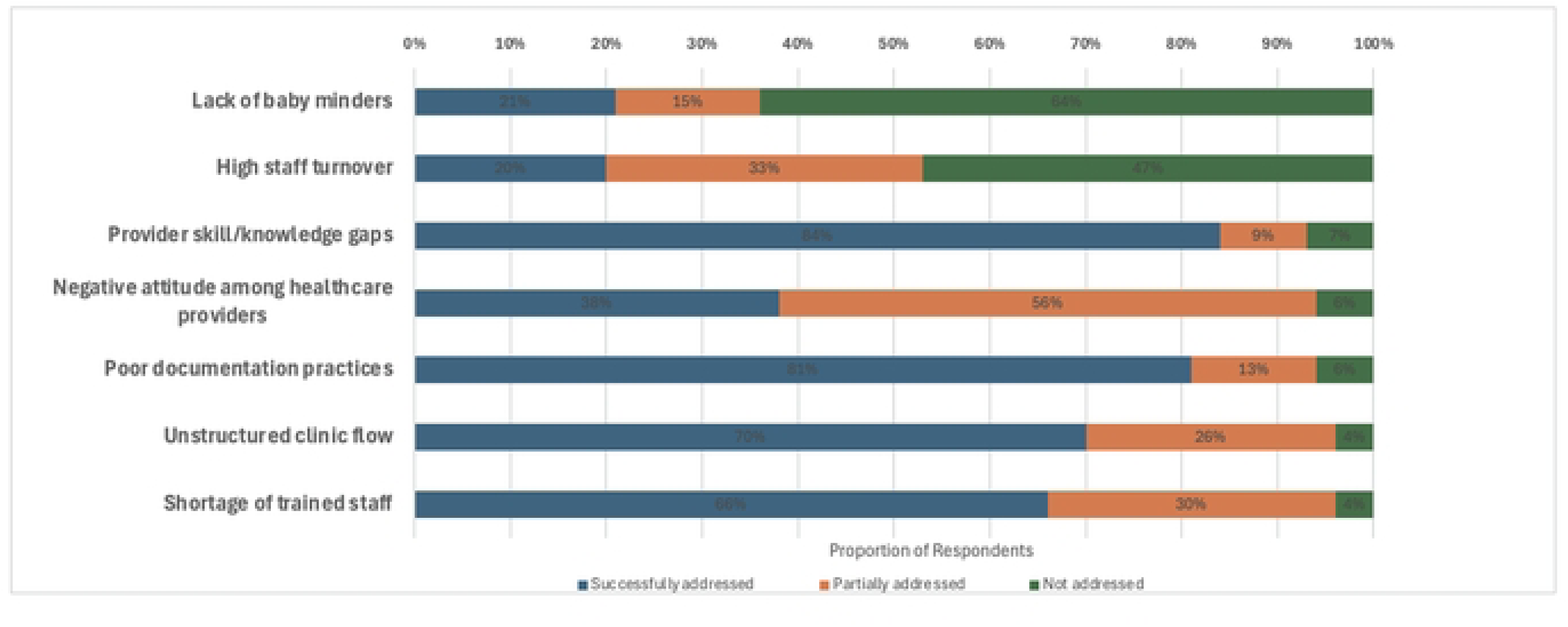
: Proportion of respondents reporting staff-related and clinic workflow barriers addressed successfully, partially, or not.

**Figure 3B.**
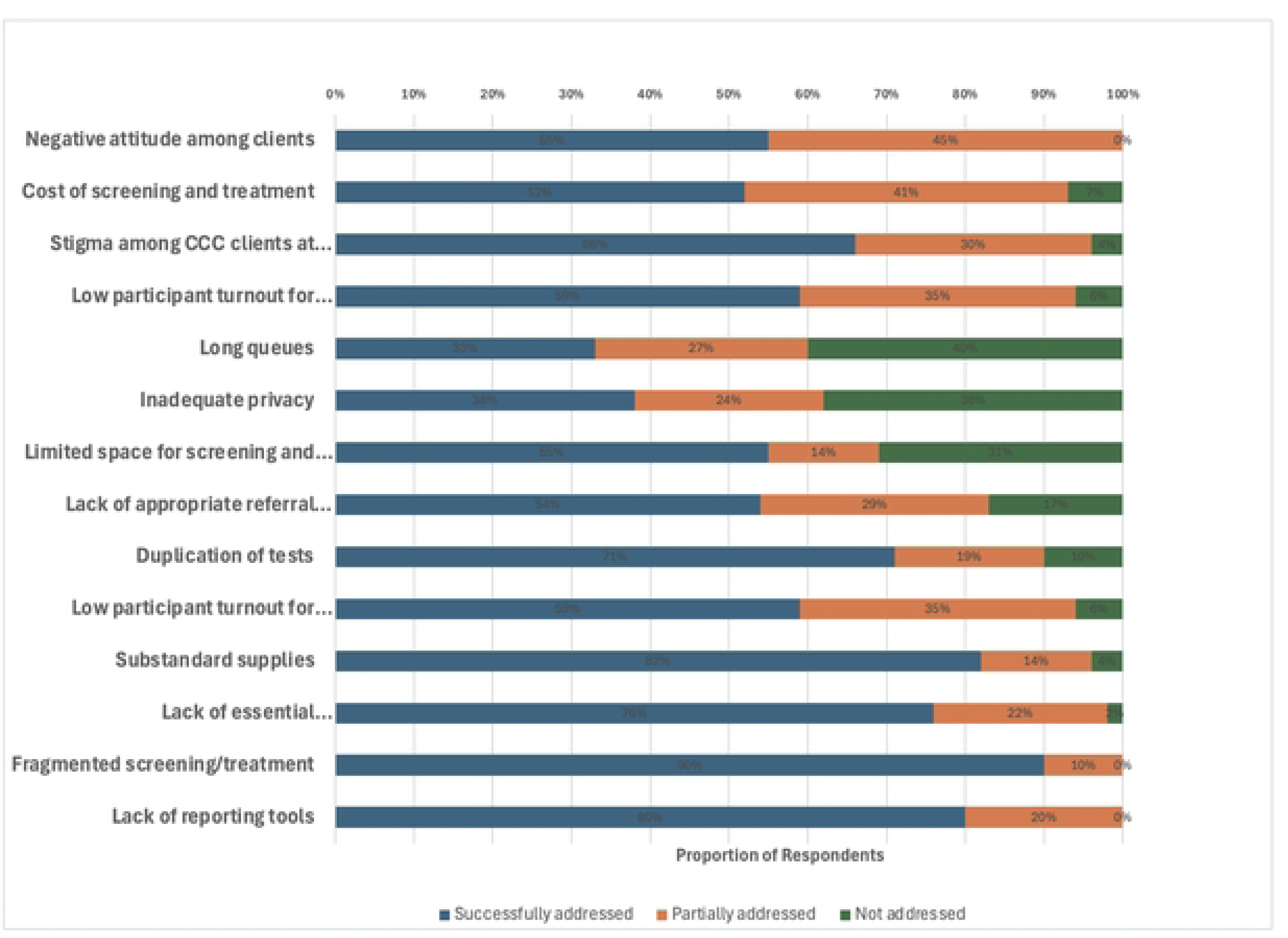
: Proportion of respondents reporting health system, infrastructure,and patient access barriers addressed successfully, partially, or not.

The barrier assessment results show that (71%, n=15) of the barriers were successfully addressed, 10% (n=2) were partially addressed, and 19% (n=4) were not addressed, as shown in Table 2 below Figures 3A and 3B categorize the barriers into two groups: (1) Staff and clinic flow barriers and (2) Health system, infrastructure, and patient access barriers, illustrating the proportion of respondents who reported barriers that were successfully, partially, or not addressed. A summary of the assessment table is also provided as supplementary material (S1 Table).

**Table 2:** Summary of barrier assessment by providers

|  | Barriers | Successfully addressed | Partially addressed | Not addressed |
| --- | --- | --- | --- | --- |
| 1 | Unstructured clinic flow (movement of clients back and forth between registration, service delivery points and cashier's office) | 16 | 6 | 1 |
| 2 | Shortage of trained staff on cervical cancer screening and treatment (SOPs), job aids, and cervical cancer screening and treatment guidelines | 25 | 11 | 1 |
| 3 | Lack of essential medication supplies and equipment. | 28 | 8 | 1 |
| 4 | Low participant turnout for screening | 20 | 12 | 2 |
| 5 | Provider knowledge gap and skills on cervical cancer screening and treatment | 27 | 3 | 2 |
| 6 | High staff turnover | 3 | 5 | 7 |
| 7 | Limited space for screening and treatment | 16 | 4 | 9 |
| 8 | Cost of screening and treatment | 14 | 11 | 2 |
| 9 | Negative attitude among healthcare providers | 7 | 10 | 1 |
| 10 | Negative attitude among clients | 17 | 14 | 0 |
| 11 | Poor documentation of cervical cancer preventive services | 13 | 2 | 1 |
| 12 | Fragmented approach to screening and treatment | 18 | 2 | 0 |
| 13 | Stigma among comprehensive care clinic clients at MCH | 25 | 11 | 1 |
| 14 | Substandard supplies especially acetic acid. (Facilities use white vinegar bought from the stores) | 18 | 3 | 1 |
| 15 | Lack of baby minders | 3 | 2 | 9 |
| 16 | Lack of participant follow-up | 10 | 14 | 4 |
| 17 | Lack of appropriate referral systems | 15 | 8 | 5 |
| 18 | Long queues | 5 | 4 | 6 |
| 19 | Inadequate privacy (screening room used for other services, students present when services are being offered) | 8 | 5 | 8 |
| 20 | Duplication of tests (VIA positive participants sent for pap smear) | 15 | 4 | 1 |
| 21 | Lack of reporting tools | 8 | 2 | 0 |

No. of barriers successfully addressed=15, partially addressed and not addressed =4.

A range of implementation strategies was applied to address the barriers reported by health providers. For example, clinic workflows were redesigned to reduce bottlenecks and improve patient flow, including the establishment of dedicated rooms for cervical cancer screening and treatment. Care coordination was strengthened by introducing patient navigators/customer care representatives to guide patients through the clinics, and health provider capacity was built through regular training sessions and continuous medical education (CME).

Additionally, some strategies addressed multiple barriers simultaneously. For example, the dual strategy of health care provider training and mentorship on cervical cancer screening and treatment helped address multiple barriers more efficiently. In this case, shortage of trained providers, provider knowledge gaps and skills on screening and treatment, use of substandard acetic acid for screening, poor documentation of cervical cancer preventive services, and duplication of screening tests were tackled with the aforementioned approach.

Strategies were mapped to the WHO health systems framework and building blocks. These strategies primarily targeted the health service delivery and health workforce building blocks, which are key health system inputs aiming at improving health and efficiency.^24^ These areas addressed key barriers such as gaps in provider knowledge, staffing shortages, and inefficiencies in clinic workflows. The mapping also highlighted target actions in essential medicine, e.g., ensuring availability of essential commodities, leadership and governance, and financing (reduction in screening and treatment costs). Additionally, community engagement emerged as a critical strategy for addressing barriers related to low screening uptake and stigma. Figures 4A and 4B provide a summary of these strategies and their subcategories.

**Figure 4A:**
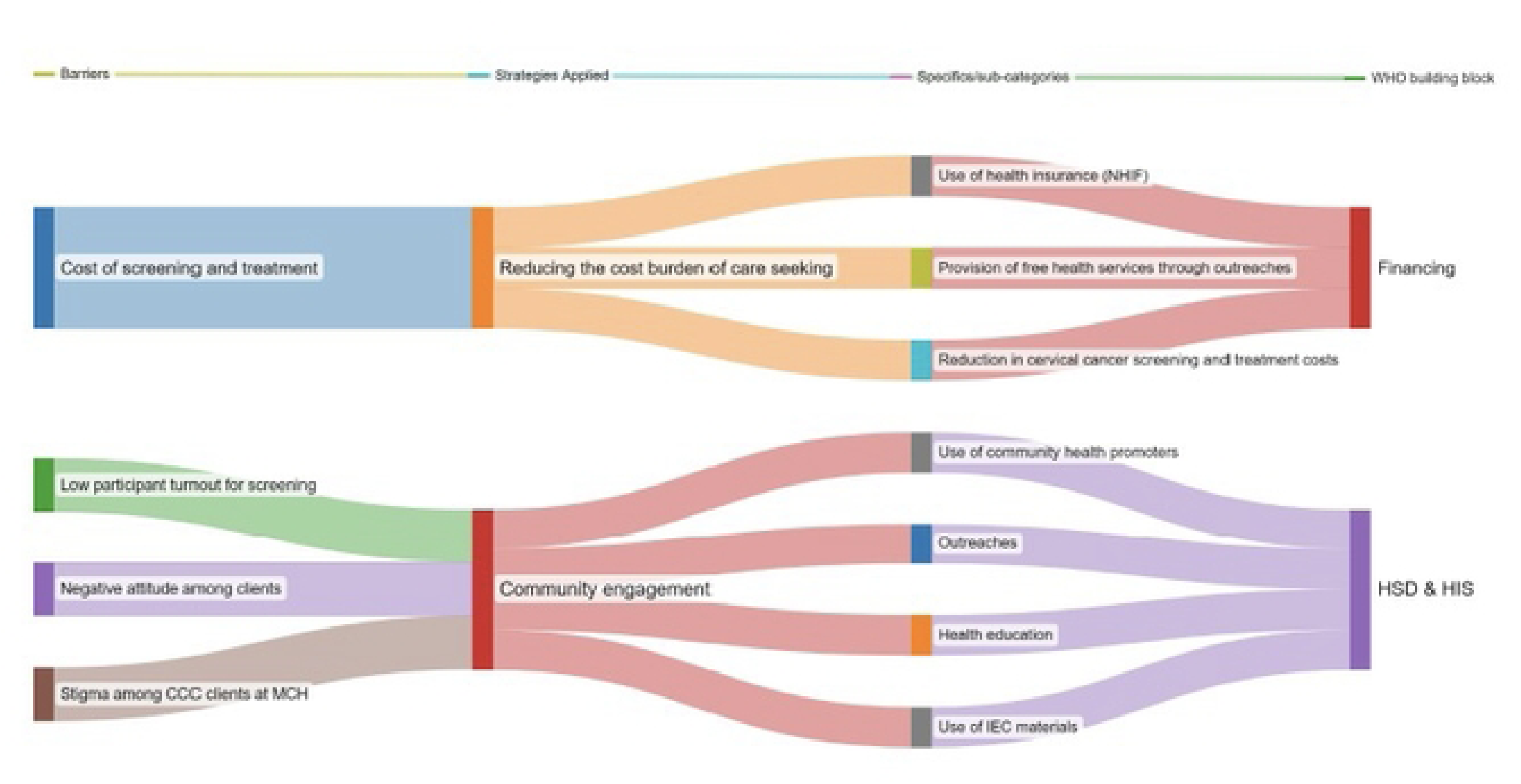
Flow diagram mappingimplementationstrategies to WHO health systems buildingblocks for addressing pre-identified patient-levelbarriers.

**Figure 4B:**
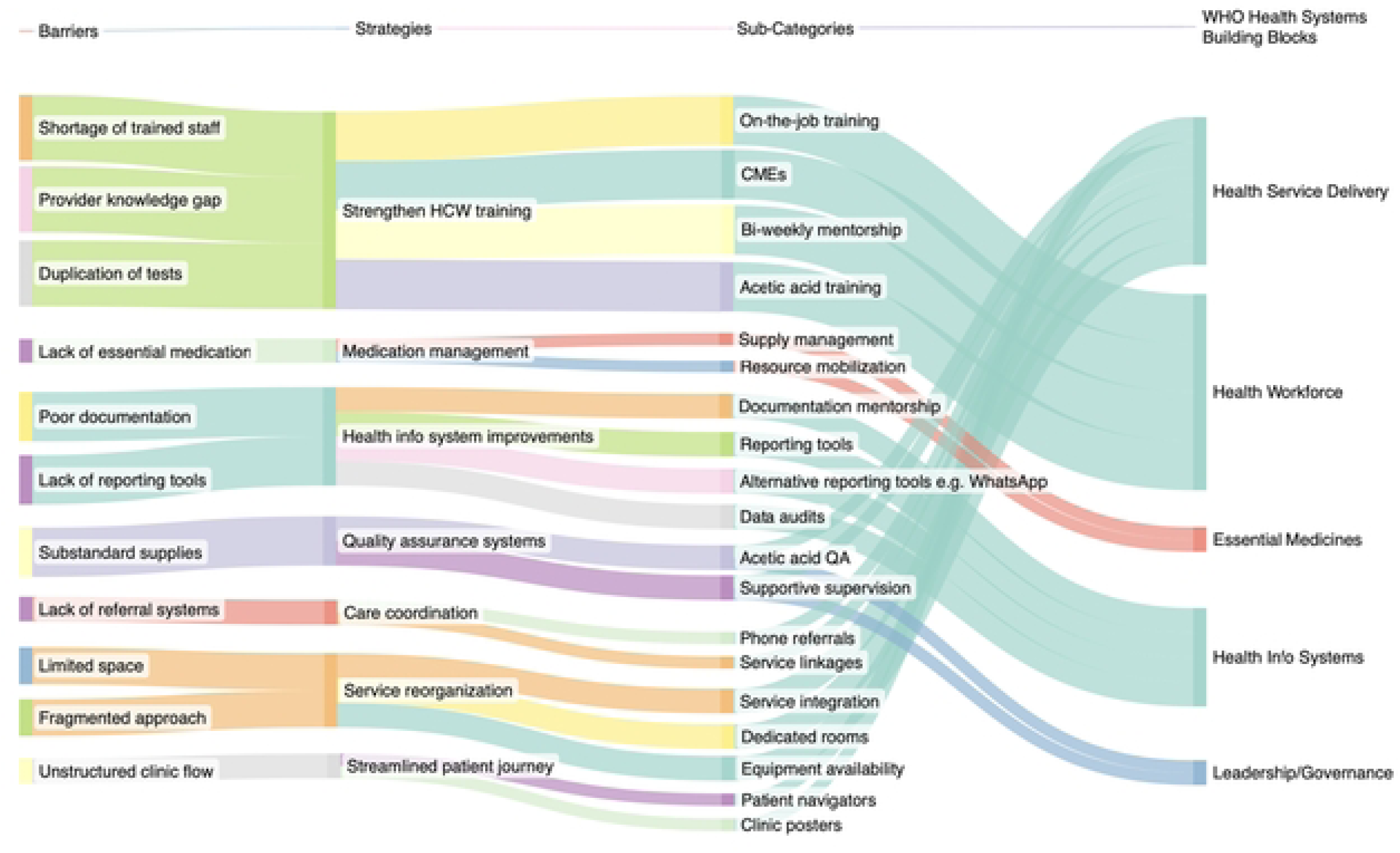
Flow diagram mapping implementation strategies to WHO health systems building blocks for addressing pre-identifiedhealth system levelbarriers.

### Qualitative Results

#### Theme 1: Improving service accessibility and optimizing clinic processes

Health care providers reported that barriers related to clinic operations, such as unstructured clinic (patient movement back and forth between registration and clinic rooms) flow, limited space for screening and treatment, fragmented approach to care, lack of essential commodities, and the cost of cervical cancer preventive services, were successfully addressed. This contributed to improved service accessibility and a more streamlined clinic process that was achieved through:

Use of customer care representatives/patient navigators to guide patients through the health system by providing direction and information on the cost and location of services offered.

> “We have a customer care lady who actually talks to patients every morning, she tells them about every section of the hospital” (Nurse, government-led facility)

a. Establishment of dedicated clinic rooms and integration of cervical cancer screening and treatment into the clinic services. The three faith-based health facilities established dedicated rooms, whereas most of the public health facilities integrated cervical cancer screening into routine clinic services such as maternal-child health and HIV clinics. However, service integration in most public health faced challenges, including delays in the provision of other services.

> *“We currently have a room for cervical cancer screening, exclusively for that purpose” (Clinical Officer, faith-based facility)*

> *“In the comprehensive care clinics (CCC), we have only one room for screening, which is also used for Operation Triple Zero (OTZ) classes. During OTZ sessions with children, screening cannot take place.” (Clinical Officer, government-led facility)*

a. Availability of essential commodities and equipment for cervical cancer screening and treatment. Eight facilities that reported a lack of essential supplies and commodities, having been fully addressed, received donations of thermal ablators and essential supplies such as gloves, acetic acid, and speculums from implementing partners. Additionally, three facilities reported leadership support, which helped ensure that the required commodities were purchased by their respective health facilities. These ensured there were no gaps in the provision of cervical cancer preventive services.

> *“The hospital management increased supplies such as cotton wool, clean gloves, and we were also able to get the lighting lamp. We also got support from the people’s Program; they were able to give us additional lighting lamps and a screening chair. " (Clinical Officer, government-led facility)*
b. Provision of free cancer screening and treatment services in some hospitals, along with reduced service charges and cost waivers. Given that the cost of cervical cancer preventive services was a challenge to the majority of the women, health facilities implemented various strategies to address this issue. Some health facilities organized free outreaches (mostly faith-based health facilities) targeting women who were unable to visit the facility due to financial constraints. Additionally, one faith-based facility reduced service charges after receiving donations of essential commodities for screening. The free services and cost waivers were applied in public health facilities.

> *"The services were made free, and even the gloves that we used to tell them to buy are now available in the hospitals," (Nurse, government-led facility)*

> *“Thanks to the free donations that we’ve been receiving, we’ve been able to offer these services at a lower cost. "(Nurse, faith-based facility)*

Although most participants reported that cost was a major barrier that had been successfully addressed, this was not the case in all the faith-based health facilities, as these sites rely on patient service charges to purchase the commodities. Additionally, women who screened positive for precancerous lesions or were suspected to have cancer and required additional tests and procedures, such as biopsy sample collection or Loop Electrosurgical Excision Procedure (LEEP) treatment, still experienced financial constraints, as these additional services were charged.

#### Theme 2: Capacity building to enhance provider knowledge, skills, and quality of cervical cancer preventive service delivery

Health provider training on screening using VIA and treatment of precancerous lesions with thermal ablation, conducted during the roll-out of TIBA across the health facilities, significantly enhanced providers’ knowledge and skills, while also improving their overall competency. This training was supplemented by additional opportunities for professional development, including monthly continuous medical education (CME) sessions, provision of job aids, technical assistance by the study, and on-the-job training for new staff in the reproductive health clinics.

> *"The KEMRI team came in and trained us and gave us support and job aids."(Nurse, government-led facility)*

> *"First of all, it was done through the initial training; besides that, there was continuous on-job mentorship.” (Laboratory Technician, faith-based facility)*

However, some facilities reported that staff shortages and lack of training among other personnel hindered service provision, despite the initial training that had been offered. This was an emerging barrier during the implementation phase.

> *“In terms of sustaining it, we don’t have a specific person stationed there to attend to mothers. The management can consider adding staff who will be stationed there so that these mothers, when they come during their MCH clinics, can be attended to”. (Nurse, faith-based facility)*

To further improve the quality of cervical cancer prevention services, mentorship was provided on the right formulation of acetic acid for screening. Most of the health facilities had previously relied on acetic acid sold in stores, which did not meet the required standard, and negatively affected the accuracy of the screening process. Training in this process addressed barriers related to a lack of provider knowledge on cervical cancer screening and treatment, as well as influenced the shortage of trained providers.

> *"Through training, we’ve learned the importance of using the right formulation of acetic acid with an expiry date that delivers results. Previously, we used 4-6% acetic acid, a garden vinegar bought from the supermarket, but now we can properly prepare and ensure the correct acetic acid concentration for screening." (Nurse, government-led facility)*

Challenges related to a lack of reporting tools and poor documentation were addressed through the provision of reporting tools, mentorship on proper documentation by the health records and information officers, and regular data audits during supportive supervision.

> *"We have had frequent supportive supervision, and the registers have been reviewed several times. The main issue was usually poor documentation, despite screening being conducted. However, most of these documentation issues have now been addressed." (Clinical Officer, government-led facility)*

#### Theme 3: Strengthening community engagement and communication for cervical cancer prevention

Health facilities that identified negative attitudes among clients and low participant turnout for screening as barriers addressed these challenges by successfully conducting routine health talks and sensitization at the clinics. This was achieved by the distribution of information and communication (IEC) materials, such as posters, and involving community health promoters to mobilize eligible women for cervical cancer screening services. Negative attitude towards screening was associated with fear of screening and receiving a positive result and lack of knowledge about cervical cancer. Additionally, outreach activities within various communities also played a key role in increasing women’s participation in the screening.

> *“Outreaches have been conducted and the number of women accessing services has increased.” (Clinical Officer, government-led facility)*

> *“Previously, these participants were unable to come to the facility for screening due to fear and other concerns. However, after the training, we were able to provide health education to help alleviate their fears and encourage participation." (Nurse, faith-based facility)*

Themes emerging from barriers partially addressed and not addressed included staffing challenges, workforce instability, and lack of appropriate infrastructure and support systems.

#### Theme 1: Staffing challenges and workforce instability

High staff turnover was mainly reported in faith-based health facilities. The transition was driven by better opportunities elsewhere. This led to the disruption in service delivery.

Unfortunately, this barrier could not be addressed as it was beyond the control of the providers. In contrast, public health facilities experienced more staff changeovers. Staff changeovers were partially addressed by minimizing the movement of trained staff on cervical cancer screening and treatment staff between departments. However, in certain circumstances, such as acute staff shortage, it was inevitable to avoid the changeovers from reproductive health clinics to other departments within the hospital.

> *"One significant challenge we still face is staff turnover. The reason is that many healthcare workers, when opportunities arise to work in public health facilities, often move to the public sector. (Medical Officer, faith-based facility)*

> *“Sometimes it is circumstances that force us to move a staff from one area to the next. It is beyond us. (Nursing manager, government-led facility)*

#### Theme 2: Lack of infrastructure and support systems for continuity of care

Appropriate support structures, such as baby-minder facilities, were reported unavailable, and as such, the barrier was not addressed. The participants also reported inadequate space in the screening rooms to set up a cot for the babies while mothers were being screened. This issue cut across both public-led and faith-based health facilities, and it was believed to discourage women with young children from accessing screening services as they were concerned about childcare and potential security risks by leaving their children with strangers at the waiting bay.

> *“When a mother is being screened and there is no one to hold their baby, and then also considering our setup is within a slum, you cannot be sure if the person holding the baby will go away with the baby. When we are doing mobilization, we encourage them to come with their neighbors or friends. (Nurse, government-led facility)*

Inconsistent participant follow-up, largely due to a lack of logistical support from the hospital management, was partially addressed. Providers often relied on their personal phones and support from implementing partners to make follow-ups, which created gaps in the continuity of care and subsequently decline in patient engagement and retention. This challenge was primarily common among women who screened positive and were not amenable to thermal ablation, or were suspicious for cancer, referred for further tests and procedures such as LEEP. Additionally, participants from one facility reported losing patient contact information within their data systems, which made follow-up efforts difficult. This challenge was yet to be resolved.

> *"There should be a phone in the screening rooms that should be used to contact these patients. When we need to follow up with clients, whether they missed a follow-up or completed treatment, we have to use our personal phones.” (Nurse, government-led facility)*

> *"There are limitations, like transport, which is not provided; you use your cash or airtime, but you are not reimbursed.” (Cytologist, government-led facility)*

Overall, the survey and interview data corresponded. The divergence observed in the qualitative responses was mainly related to emerging barriers rather than the pre-identified ones. These emerging barriers were not addressed, impacting TIBA implementation. For example, women with precancerous lesions covering more than 75% of the cervix and not eligible for thermal ablation did not receive timely LEEP treatment. This delay was attributed to both a shortage of gynecologists trained in LEEP and reluctance among those trained to perform the procedure. The reluctance stemmed from differing opinions on management approaches, as many gynecologists preferred performing LEEP only for patients with confirmed positive findings on pap smear or colpobiopsy, rather than solely relying on VIA results. Table 4 provides a summary of these emerging barriers.

**Table 4:** Emerging barriers during TIBA implementation

| Barrier | Description |
| --- | --- |
| Inconsistent supply of alternative screening modalities across health facilities | Alternative tests such as HPV and pap smear testing kits were inconsistently available. |
| Staff changeovers | Transfer of staff trained in cervical cancer screening and treatment to other departments hindered continuity in cervical cancer preventive services. |
| Incomplete treatment for women with larger cervical precancerous lesions | Women with larger precancerous lesions greater than 75% not amenable to thermal ablation did |
|  | not receive treatment as gynecologists were reluctant to perform LEEP |
| Cost of referral procedures such as LEEP | High costs associated with procedures like LEEP created barriers among women who screened positive for precancer to access the service. |
| Gaps in provider skills and expertise | Some healthcare providers had challenges in differentiating positive lesions from conditions such as cervicitis and ectopy. |
| Staff capacity strain due to high participant volumes during outreach services | Staff faced challenges in managing high participant turnouts during outreaches due to shortage of providers, which impacted service delivery. |

### Match between barriers and strategies

The implementation strategies deployed during TIBA implementation matched the barriers identified by the key stakeholders (healthcare providers, health facility managers, and policymakers) during the stakeholders’ workshop. As a result, these strategies successfully addressed 71% of the pre-identified barriers, with greater success in improving accessibility to cervical cancer screening and precancerous lesion treatment and enhancing provider skills. It is also important to note that there were strategy adaptations that occurred during the implementation phase. However, these adaptations are detailed in a separate paper.

## Discussion

We assessed whether the implementation strategies deployed were aligned with the pre-identified barriers to implementing TIBA. The majority (81%) of the reported pre-identified barriers were health system-related. Overall, the implementation strategies were well matched to the barriers, as most (71%) were successfully addressed. Several strategies were applied to address these barriers, with some addressing multiple barriers simultaneously. Key themes related to successful barrier-strategy matching included targeted improvements in service accessibility, streamlining and optimizing clinic processes, investing in health provider capacity building through training, and strengthening community engagement and communication. Approximately 10% of the barriers were partially addressed, while 19% were not addressed. The barriers partially addressed and those not addressed were primarily associated with structural problems, including staffing challenges, workforce instability, infrastructural limitations, and lack of support systems for participant follow-up. These issues require long-term investments and system-level changes, beyond the health providers’ control. For example, while training can improve provider skills, staff shortages and high turnover cannot be resolved immediately, as the process of recruiting new staff involves lengthy administrative processes, which are also influenced by other factors.

In this study, the pre-identified barriers to implementing TIBA, including shortage of trained providers, lack of essential commodities, poor documentation, among others (a full list of the 21 barriers is provided as S2 Table), are not unique to this context. These challenges have been widely reported in similar settings implementing cervical cancer prevention services in countries like Nigeria, Zambia, Malawi, Rwanda, and counties in Latin America.^25–28^ To address these barriers, several implementation strategies, also highlighted in the literature, were employed, including provider training and mentorship, availability and management of essential commodities, supplies, and equipment, supportive supervision, community engagement through routine health education, and community outreaches.^29^ We mapped these strategies to the WHO health systems framework, and most strategies primarily targeted health service delivery, while others addressed key health system inputs such as health workforce and access to essential commodities, aiming at improving overall health system performance and efficiency.^24^ These findings suggest that a “one-size-fits-all” approach may not be sufficient in addressing contextual challenges.^30^ For instance, in this study, some faith-based health facilities addressed the barrier of cost by reducing charges when they received donations of screening commodities or by offering free outreaches. In contrast, some public-led health facilities provided free routine screening and treatment services along with cost waivers for additional tests required.

Notably, some strategies addressed multiple barriers simultaneously. For example, provider training and mentorship on cervical cancer screening and treatment addressed provider knowledge gaps and skills, reduced the shortage of trained providers, improved documentation, ensured use of standard/required acetic acid formulation, and minimized duplication of screening tests. These findings have two key implications. First, they suggest that some implementation strategies, such as improving health worker skills, can have a ripple effect in improving several areas of care, including screening quality, treatment consistency, and reliability of care.^31,32^ Second, it underscores the interconnected nature of barriers, highlighting the need for a holistic approach in making adaptable changes in health service delivery, particularly in resource-constrained settings, to optimize the use of available resources.^33,34^

It is important to note that, while most of the health providers reported having successfully addressed challenges related to the availability of cervical cancer essential commodities and equipment, these were largely addressed with support from implementing partners.

Considering the recent USA funding cuts, which have impacted several vertical programs in LMICs, including cervical cancer prevention efforts,^35^ have led to delays in screening and precancer treatment. These delays could potentially lead to devastating outcomes, hindering global efforts towards cervical cancer elimination. To build resilient and sustainable health systems, LMICs must prioritize the development and implementation of national plans for cervical cancer prevention. This can be achieved through domestic resource mobilization, strengthening planning and prioritization processes, and gradually transitioning away from reliance on volatile donor assistance.^36,37^

Pre-identified barriers, partially addressed or not addressed, were related to health systems gaps beyond the health provider’s control. These included infrastructural challenges such as a lack of baby minder spaces due to space limitations, high staff turnover, especially in faith-based health facilities, and a lack of participant follow-up due to logistical and data system challenges. These barriers, common in other LMICs,^38,39^ derail progress toward cervical cancer elimination by creating missed opportunities for timely interventions and leading to inefficient resource use, especially when trained providers are relocated or transferred to other departments or health facilities.^40^ Addressing these challenges through improved infrastructure, better staff retention strategies and enhanced participant follow-up is crucial for advancing cervical cancer prevention.^41^

Additional barriers emerged during the implementation of TIBA. These included incomplete treatment for screen-positive women not eligible for treatment with thermal ablation, often due to cost constraints or reluctance from health providers (especially gynecologists).

Challenges also arose in differentiating positive lesions from other cervical conditions, such as cervicitis and ectopy, particularly among newer staff who had not undergone initial training despite technical assistance provided by the study team during the intensive implementation phase. While initial VIA trainings and technical assistance offered by implementing partners can support early implementation, they are often short-term. Without structured and sustained opportunities for retraining, ongoing mentorship, and supportive supervision created by the health management teams, there is a clear risk of declining diagnostic skills among providers.^42^ Therefore, alongside continued in-person mentorship, integrating VIA mentorship through telemedicine platforms could offer a viable option to improve the quality of screening and expand service reach.^42,43^

Screening alone is ineffective without timely and appropriate linkage to treatment.^41,44^ In this study, the single-visit, screen-and-treat approach was not feasible for women with lesions more than 75% and extending to the ectocervix, as they required LEEP treatment. However, access to LEEP was hindered by several factors, including, lack of LEEP equipment, shortage of trained providers on LEEP and out-of-pocket costs was a limitation to most women. These findings have been documented,^45,46^ and our study additionally highlights that some gynecologists were reluctant to perform LEEP for women who screened positive for VIA, despite national guideline recommendations for precancer treatment.^47^ Instead, they preferred performing LEEP on patients with positive findings on pap smear or colpobiopsy.

Non-adherence to guidelines has been associated with delays in treatment, disease progression, and an increase in health expenditure.^48,49^ To strengthen cervical cancer prevention efforts, there is an urgent need to expand the pool of trained LEEP providers and implement task sharing for LEEP services with ongoing supervision and mentorship.

Additionally, quality improvement interventions are needed to ensure guideline adherence and timely access to treatment for screen-positive women.

A key strength of this study is its mixed methods design, which allowed integration of quantitative data to examine barrier-strategy matching, alongside qualitative insights that provided a deeper understanding of drivers behind successful, partial, and unsuccessful strategy implementation. This comprehensive approach enhanced the validity of our findings. However, the study should be interpreted considering some limitations. Of the ten health care facilities included, survey data and participant perspectives were obtained from only two health facility managers. As a result, we may have missed certain insights from a broader management perspective that could have influenced strategy implementation.

Additionally, this study was designed to obtain data primarily from health care providers. Including perspectives from women who received cervical cancer prevention services could have provided more insights into the extent to which patient-level barriers were matched across these health facilities, as well as contextual factors that influenced their experiences.

## Conclusion

In this study, the majority of the pre-identified barriers were successfully matched to the deployed implementation strategies, which mainly targeted improving health service delivery, workforce capacity, and community engagement. Applying the appropriate strategy to address barriers is associated with a higher probability of achieving the desired outcomes and maximization of the intervention’s impact. Despite the overall success, important gaps remain, particularly related to structural challenges, such as staffing instability, infrastructural limitations, gaps in provider skills, and lack of support systems for continued care in cervical cancer prevention. These findings highlight the need for a more adaptable, context-specific strategies and sustained investments in health systems strengthening, with minimal reliance on external donor support, to achieve resilient and effective cervical cancer prevention programs in resource-constrained settings.

### Implications

- Multiple strategies were applied to address barriers across health facilities. Although some strategies were similar, their application varied depending on context, for example, faith-based vs public health facilities. Adaptations to context enhance strategy effectiveness. Hence, flexibility and context-specific implementation are critical for success.
- Some implementation strategies addressed multiple barriers simultaneously. The interconnected nature of barriers highlights the need for a holistic approach, which is key in optimizing resource use, particularly in resource-constrained settings. Integrated strategies may be more impactful in accelerating implementation success than isolated ones.
- Essential commodity management and equipment availability (thermal ablators) were largely supported by implementing partners.

With donor cuts, we are experiencing service disruptions like treatment delays. Strengthening sustainable health systems through local ownership is critical to building resilient and self-reliant cervical cancer prevention services.

- Emerging provider skills gap in screening using VIA.

Initial trainings are short-term (for example, VIA) and there is a risk of declining diagnostic skills if not followed by a structured retraining and sustained mentorship.

- Screening is ineffective without timely treatment.

Delays in receiving LEEP treatment were reported due to equipment shortages, few trained providers and out-of-pocket costs.

Gynecologists’ reluctance and non-adherence to guidelines further delayed treatment.

There is a need to expand the pool of LEEP-trained providers, implement task sharing, and strengthen quality improvement and supervision.

## Data Availability

Data can be accessed at https://figshare.com/s/924521187981b129920d

